# Validation of the International IgA risk prediction tool in American Indians and Hispanics

**DOI:** 10.1101/2023.06.29.23292045

**Authors:** Saeed Kamran Shaffi, Edger Fischer, Christos Argyropoulos, Brent Wagner

**Affiliations:** Staff Nephrologist, Raymond G. Murphy VA Medical Center Associate Professor, Department of Internal Medicine, Division of Nephrology, University of New Mexico; Professor of Pathology Division of Surgical Pathology and Cytopathology University of New Mexico; Associate Professor, Department of Internal Medicine, Division of Nephrology, University of New Mexico; Director, Kidney Institute of New Mexico University of New Mexico Health Sciences Center ACOS, Research Service New Mexico VA Health Care System*

**Keywords:** Immunoglobulin A Risk Prediction, Risk Prediction Model Calibration Assessment, Risk Prediction Model Discrimination Assessment

## Abstract

**Background:** The International Immunoglobulin A nephropathy (IgAN) risk prediction assesses the risk of kidney failure in patients with IgAN. The performance of this risk prediction tool has not been studied in American Indians and Hispanics. We conducted a single-center study to assess the equation performance in this population.

**Methods:** We calculated the 5-year risk of developing kidney failure with the IgAN risk prediction equation without race and assessed the equation performance using the metrics of calibration, discrimination, and overall prediction error.

**Results:** Thirty-four patients were included, most of whom identified as of Hispanic race/ethnicity (44.1%), or as American Indians (26.5%). At biopsy, the median (IQR) age, serum creatinine, and spot urine protein to creatinine ratio were 38 years (27-45), 2.15 mg/dl (1.51-3.04), and 2.7 g/g (1.5-5.8), respectively. The equation identified patients at high risk of developing kidney failure early with a concordance statistic of 0.79 (95% CI 0.68 – 0.89). The agreement between observed and predicted outcomes at 5 years was marginal, with over-estimation of risk for patients with low observed risk and vice versa. Overall prediction error was suboptimal in this cohort [index of prediction accuracy 0.34 (0.03 – 0.51)].

**Conclusions:** The International IgAN risk prediction equation without race accurately identified patients at elevated risk of developing kidney failure. At 5 years, the agreement between the observed and predicted outcomes was sub-optimal, possibly due to advanced kidney disease in this cohort. A diverse development population may improve the risk prediction.

## Introduction

IgA nephropathy (IgAN) is one of the most common primary glomerular diseases worldwide, with an incidence of 2.5 per 100,000 in adults. It is associated with end-stage kidney disease (ESKD) in 10-40% of patients by ten years of diagnosis [1]. Our understanding of IgAN pathophysiology has improved due to recent advances, which have led to new therapies [2, 3]. Therefore, it is imperative to identify patients who would rapidly progress to kidney failure and would benefit from early goal-directed treatments. Furthermore, an accurate risk prediction tool - that identifies patients at an elevated risk of developing kidney failure - would assist patients and clinicians to prepare for transplant and renal replacement therapy and instituting measures to decrease cardiovascular risk, which is associated with mortality in patients with chronic kidney disease [4].

To address these issues, Barbour et al. developed an International IgA risk prediction tool that uses demographic, clinical, and kidney biopsy variables to assess the risk of kidney failure or >50% decline in the estimated glomerular filtration rate (eGFR) [5]. The authors developed two equations with and without an ethnic variable (Chinese, Japanese, or Other). The equation without a race/ethnicity variable was developed (n = 2781) and validated (n = 1146) in large cohorts comprising Europeans, Chinese, and Japanese. Since the publication of the original report, IgAN risk prediction tool performance has been studied in Chinese [6, 7], South Asian [8], Greek [9], and South American [7] cohorts. Table 2 summarizes the performance of the equation without the race/ethnicity variable in these cohorts. The equation performance has not been studied in the Hispanic and American Indian populations. A New Mexico based kidney biopsy registry data analysis has revealed a higher frequency of IgAN in the American Indians compared to their state population share which may point towards a higher incidence of IgAN in them [10]. Therefore, we conducted this single-center study to assess the performance of the International IgA risk prediction tool without race in a cohort from New Mexico where, Hispanics and American Indians are in the majority.

## Methods

The University of New Mexico Kidney Biopsy registry was established by searching for the current procedural terminology (CPT) codes for kidney biopsies performed at the University of New Mexico (UNM) hospital between 2001 and 2016. The institutional review board at UNM approved the Kidney Biopsy Registry. The principal investigator (SKS) created a HIPPA complaint online Kidney Biopsy registry. We queried the complete biopsy report for the following terms: “Immunoglobulin A nephropathy,” “IgA Nephropathy,” and “IgAN.” We reviewed the biopsies to identify patients with primary IgAN who met the inclusion criteria. The IgA risk prediction tool without the race variable uses demographic and clinical variables at the time of biopsy that includes chronic kidney disease epidemiology (CKD-EPI) equation estimated GFR, mean arterial blood pressure, proteinuria, age, the Oxford kidney biopsy classification scores of mesangial hypercellularity, endothelial proliferation, segmental sclerosis, and tubular atrophy, and medication use of angiotensin-converting enzyme inhibitor or angiotensin receptor blocker, and immunosuppression around the biopsy to predict the risk of kidney failure defined as: 1) end-stage kidney disease requiring dialysis or a kidney transplant or an estimated glomerular filtration rate of < 15 ml/min/1.73m^2^ – or, 2) >50% decline in eGFR from the baseline value. For this study, we included patients with primary IgAN and on whom the International IgAN Prediction Tool without race variable predictors were available. Other inclusion criteria included the availability of longitudinal data for at least 12 months. The patients who developed kidney failure or had >50% decline in the eGFR within 12 months were also included. We excluded patients who were < 18 years of age, who had kidney failure at the time of biopsy, and on whom the risk prediction variables or longitudinal laboratory data to assess risk were unavailable. The primary outcome was the development of end-stage kidney disease or an eGFR <15 ml/min/1.73m^2^ or a > 50% decline in the estimated GFR from the value at the time of biopsy.

We summarized the baseline variables stratified on race and ethnicity and reported the categorical variables as n (%) and continuous variables as median (IQR). For comparisons between groups, we performed the Kruskal-Wallis Rank Sum test or Fisher’s Exact test as appropriate. A renal pathologist (EF) reviewed all the biopsies and assigned them a MEST-C score. For survival analysis, patients were censored when they were lost to follow-up or at the end of the study.

We calculated the linear predictors and the predicted probability of the primary outcome using the formula without the race variable (S1) as we had few patients with the Chinese or Japanese race in our cohort.

We assessed the performance of the equation in our cohort using the metrics of calibration, discrimination, and the overall prediction error [11, 12]. A well-calibrated risk assessment model’s predictions should match the observed outcomes. Ideally, a model’s predictions should not only match the observed effects at a particular time (e.g., 5 years) - known as calibration in the large - as well as across the entire range of predictions. The population is usually divided into risk categories for calibration assessment. The mean observed risk for each category is obtained from a Kaplan-Meyer model and risk predictions from the risk equation. For each category, the observed and predicted risks are plotted to assess agreement. There are a few issues with this approach; first, the risk categories are arbitrary and result in loss of information, and second, this method cannot account for censored patients. To circumvent these issues, we used the pseudo-observations approach for the primary outcome indicator as recommended by the Strengthening Analytical Thinking of Observations Studies (STRATOS) initiative[11, 12]. Observed risk calculations employed weighting techniques with pseudo-observations accounting for the censored patients. We regressed the primary outcome with the linear predictor by performing a Cox-proportional Hazard analysis. We obtained the 5-year estimated and observed risk and plotted them to obtain a calibration plot. The calibration plot should follow a 45-degree line for a well-calibrated equation; risk estimate values above and below the 45-degree line suggest under and over-prediction, respectively. We also obtained numerical summaries of calibration. The ratio of the observed and expected outcomes (O/E) should be 1; a ratio less than 1 signifies overprediction and greater than 1 under-prediction. We obtained the O/E ratio at 5 years. We regressed the primary outcome with the log-transformed predicted risk using a restricted cubic splines model with smoothing to obtain the calibration intercept and slope. An intercept of 0 means that, on average, the observed and the predicted risks are well matched. A slope greater than or less than 1 indicates the homogeneity and heterogeneity of the risk estimates, respectively.

Discrimination assesses the ability of a risk model to identify patients who had the outcome of interest earlier than others. We regressed the primary outcome with the linear predictor to assess discrimination and calculated the concordance statistics (C-Statistic). The area under the curve (AUC) was graphically represented by plotting the true positive rate (sensitivity) against the false positive rate (1 – specificity).

The index of prediction accuracy (IPA) summarizes both calibration and discrimination and represents the overall prediction error [13]. We used the following formula to calculate IPA:

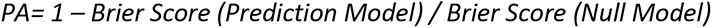

Where the Brier score is the mean squared difference between the primary outcome indicator and risk estimates, the prediction model is a Cox-proportional Hazards model regressed with the linear predictor, and the null model had no predictors. A higher IPA score indicates better model performance.

We used bootstrapping approach (100 bootstraps with data replacement) for estimating the area under the curve, C-statistics, and IPA to ensure that the standard errors and confidence intervals for these metrics were accurate.

## Results

We identified 74 patients with suspected IgA nephropathy by searching the University of New Mexico Kidney biopsy registry (Fig 1). Out of 74, 34 patients met the eligibility criteria. The median age at the time of the kidney biopsy was 38 years (IQR 27-45 years) [Table 1]. Most of the patients identified as of the White race (38%). Of the patients who identified as White, the majority were of Hispanic ethnicity (53%). We collated race and ethnicity into a single variable and observed that about 3 out of 7 patients were of Hispanic race/ethnicity (44.1%), 2 out of 7 identified as of American Indian race/non-Hispanic ethnicity (26.5%), and the rest were of other races and ethnicities (29.4%). The other category included Asians (n = 3), non-Hispanic Whites (n=3), African Americans (n =1), and of different races or ethnicities or unknown races and ethnicities (n=3).

**Figure 1:**
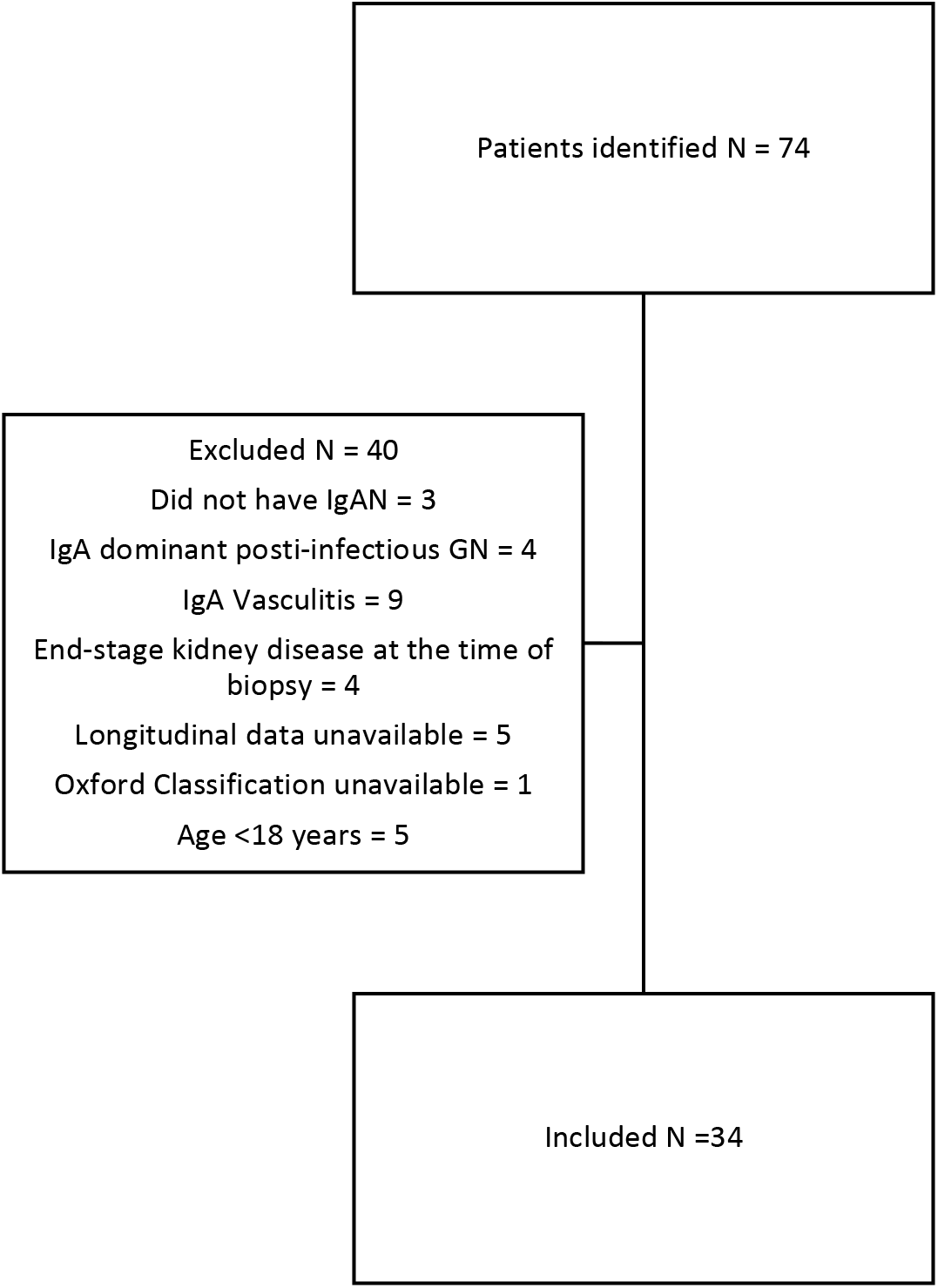
Flow diagram showing the patients identified, excluded, and included in the study

**Table 1:** Demographic, Clinical, and the Oxford Classification of the patients with IgAN stratified on Race and Ethnicity

|  | Overall, N = 34<sup>‡</sup> | Hispanic, N = 15<sup>‡</sup> | American Indian, N = 9<sup>‡</sup> | Other, N = 10<sup>‡</sup> | p-value<sup>‡</sup> |
| --- | --- | --- | --- | --- | --- |
| Age; Median (IQR) | 38 (27 - 45) | 35 (29 - 47) | 40 (26 - 44) | 40 (25 - 50) | >0.9 |
| Sex; n(%) |  |  |  |  | >0.9 |
| Female | 18 (53%) | 8 (53%) | 5 (56%) | 5 (50%) |  |
| Male | 16 (47%) | 7 (47%) | 4 (44%) | 5 (50%) |  |
| Race; n(%) |  |  |  |  | <0.001 |
| White | 11 (32%) | 8 (53%) | 0 (0%) | 3 (30%) |  |
| American Indian | 9 (26%) | 0 (0%) | 9 (100%) | 0 (0%) |  |
| Unknown | 6 (18%) | 4 (27%) | 0 (0%) | 2 (20%) |  |
| Asian | 3 (8.8%) | 0 (0%) | 0 (0%) | 3 (30%) |  |
| Hispanic | 3 (8.8%) | 3 (20%) | 0 (0%) | 0 (0%) |  |
| Other | 1 (2.9%) | 0 (0%) | 0 (0%) | 1 (10%) |  |
| Black/African American | 1 (2.9%) | 0 (0%) | 0 (0%) | 1 (10%) |  |
| Creatinine (mg/dl); Median (IQR) | 2.15 (1.51 - 3.04) | 2.10 (1.56 - 3.03) | 2.41 (1.80 - 3.99) | 1.70 (1.40 - 2.75) | 0.4 |
| eGFR (ml/min/1.73m<sup>2</sup>); Median (IQR) | 34 (20 - 51) | 34 (22 - 48) | 27 (11 - 41) | 40 (31 - 56) | 0.4 |
| Baseline Proteinuria (g/g); Median (IQR) | 2.7 (1.5 - 5.8) | 3.6 (1.6 - 6.1) | 3.6 (2.0 - 4.3) | 1.6 (1.1 - 2.7) | 0.3 |
| Mean Arterial BP (mmHg); Median (IQR) | 99 (89 - 104) | 97 (90 - 107) | 100 (97 - 103) | 95 (85 - 103) | 0.5 |
| Immunosuppression around biopsy; n (%) | 12 (35%) | 6 (40%) | 6 (67%) | 0 (0%) | 0.006 |
| ACEi/ARB around biopsy; n (%) | 21 (62%) | 9 (60%) | 6 (67%) | 6 (60%) | >0.9 |
| Primary Outcome; n (%) | 23 (68%) | 12 (80%) | 6 (67%) | 5 (50%) | 0.3 |
| M; n (%) |  |  |  |  | 0.8 |
| M0 | 10 (29%) | 5 (33%) | 3 (33%) | 2 (20%) |  |
| M1 | 24 (71%) | 10 (67%) | 6 (67%) | 8 (80%) |  |
| E; n (%) |  |  |  |  | >0.9 |
| E0 | 20 (59%) | 9 (60%) | 5 (56%) | 6 (60%) |  |
| E1 | 14 (41%) | 6 (40%) | 4 (44%) | 4 (40%) |  |
| S; n (%) |  |  |  |  | 0.049 |
| S0 | 4 (12%) | 0 (0%) | 1 (11%) | 3 (30%) |  |
| S1 | 30 (88%) | 15 (100%) | 8 (89%) | 7 (70%) |  |
| T; n (%) |  |  |  |  | 0.5 |
| T0 | 1 (2.9%) | 1 (6.7%) | 0 (0%) | 0 (0%) |  |
| T1 | 24 (71%) | 9 (60%) | 6 (67%) | 9 (90%) |  |
| T2 | 9 (26%) | 5 (33%) | 3 (33%) | 1 (10%) |  |
| C; n (%) |  |  |  |  | 0.8 |
| C0 | 22 (65%) | 10 (67%) | 5 (56%) | 7 (70%) |  |
| C1 | 12 (35%) | 5 (33%) | 4 (44%) | 3 (30%) |  |

<sup>‡</sup> Median (IQR); n (%)<sup>‡</sup> Kruskal-Wallis rank sum test; Fisher's exact test

At the time of biopsy, the median (IQR) serum creatinine and Chronic Kidney Disease Epidemiology equation (CKD-EPI) eGFR were 2.15 mg/dl (1.51 – 3.04) and 34 ml/min/1.73m^2^ (20-51), respectively (Table 1). Median (IQR) proteinuria assessed by a spot urine protein to creatinine ratio around the time of biopsy was 2.7 g/g (1.5-5.8). Mean arterial blood pressure was 99 mm Hg (IQR 89 -104 mm Hg). Thirty-five percent of the patients were on immunosuppression, and 62% were either on an angiotensin-converting enzyme inhibitor or an angiotensin receptor blocker within 12 months of IgAN diagnosis. American Indians showed a non-significant trend towards a higher serum creatinine, mean arterial pressure, and a lower CKD-EPI estimated GFR than the other races. American Indians and Hispanics showed a trend towards higher proteinuria around the time of biopsy than the other races, but it was not statistically significant. On the Oxford classification of IgAN, Hispanics were more likely (100%) to have an S1 score than the other races (p 0.049). Of note, 89% of the American Indians had an S1 score on kidney biopsy.

Figure 3 shows the plot of the 5-year estimated and observed risk. The observed risk was lower for the estimated risks between 20 and 50%; therefore, the model overestimated risk. The observed risk was higher for the estimated risks above 50%, indicating an underestimation of risk by the model; however, due to a small sample size, the confidence intervals were wide indicating imprecision of our findings.

Table 3 summarizes the numerical summaries of model calibration. At five years, the observed and estimated risk ratio (O/E) was less than 1 [0.88 (95% CI (Confidence Interval) 0.51-1.26)], indicating over-prediction; however, the confidence intervals crossed 1. The intercept was close to 0 [0.10 (CI – 0.37 - 0.58)]; since the confidence intervals crossed 0, we cannot make a conclusive statement about the intercept.

**Table 2:**
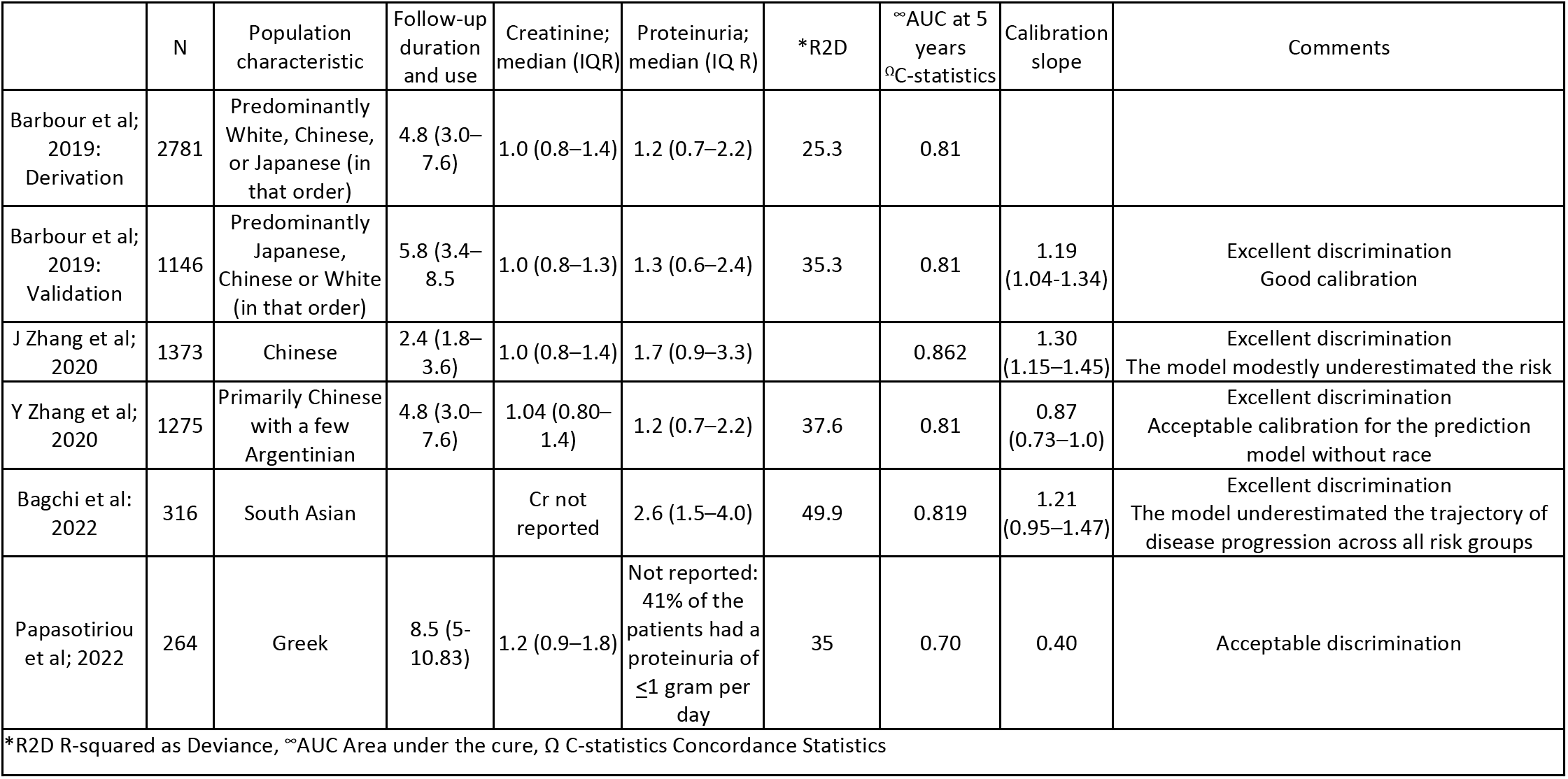
Summary of the studies assessing the performance of the international IgAN risk prediction tool in various populations

**Table 3:** Numerical summary of the measures of the International IgAN Risk Prediction Tool without race performance in our cohort

327Table 3: Numerical summary of the measures of the International IgAN Risk Prediction Tool without race 328performance in our cohort
| **Discrimination** |  |
| --- | --- |
| Metric | Value |
| C-Statistics at 5 years | 0.79 (0.64 – 0.91) |
| Area Under the Curve at 5 years | 0.79 (0.68 – 0.89) |
| **Calibration** |  |
| Observed/Estimated Risk at 5 years (O/E) | 0.88 (0.51 – 1.25) |
| Calibration Intercept | 0.10 (-0.37 – 0.58) |
| Calibration Slope | 3.03 (0.43 – 5.64) |
| **Overall Prediction Error** |  |
| Index of Prediction Accuracy (IPA) | 0.33 (0.02 -0.50) |

The risk prediction equation identified patients at a greater risk of developing the primary outcome early with a C-statistic of 0.79 (0.68 - 0.89). Figure 2 shows the receiver operator curve (ROC) with the area under the curve, which was 0.78 (0.58 -0.98).

**Figure 2:**
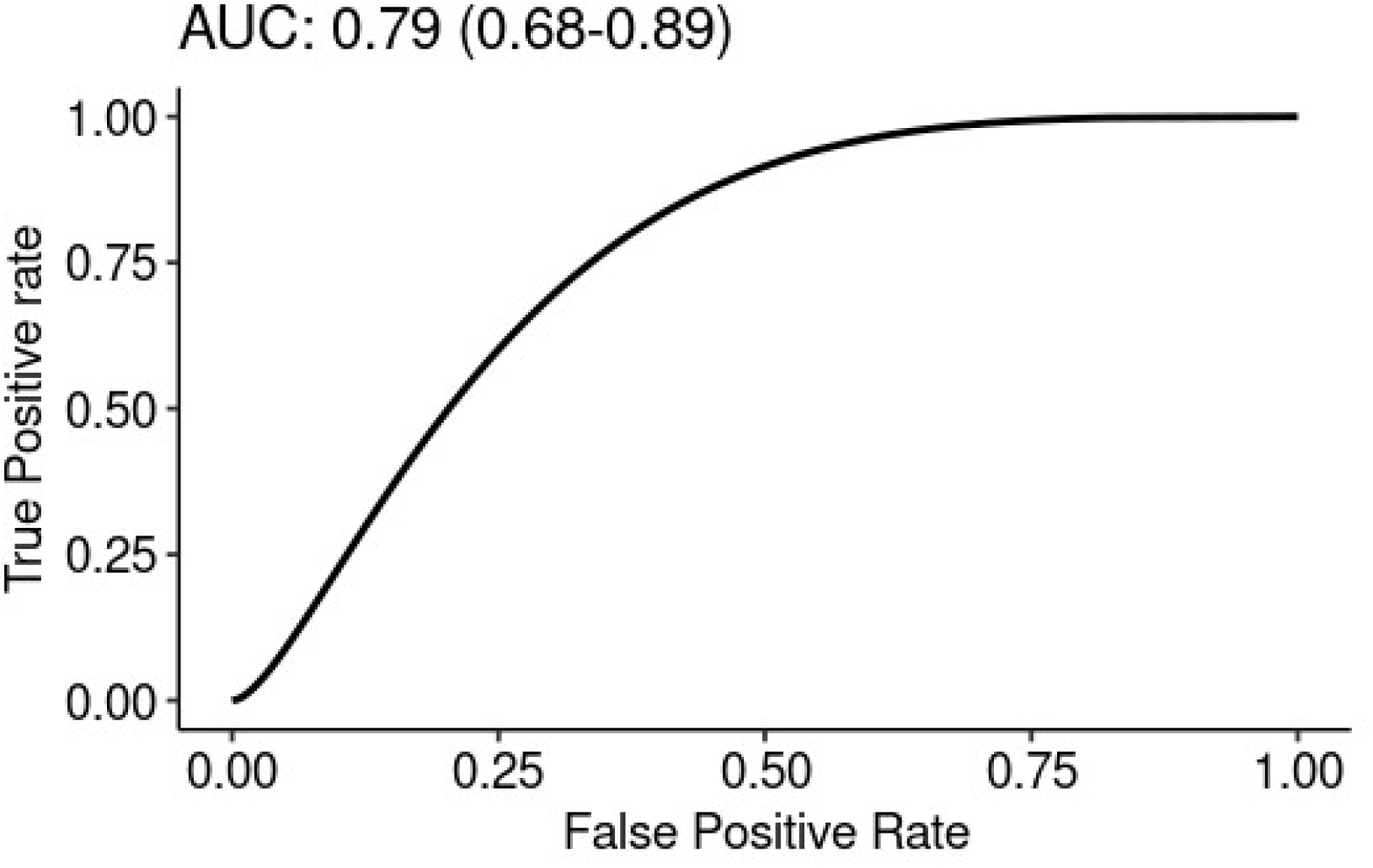
Receiver operator curve (ROC) at 5-years of the International IgAN risk prediction tool in the study population.

The prediction accuracy index was 0.34 (0.03 -0.51) (table 3), which was marginal due to poor model calibration in our population.

In summary, in our cohort, the international IgA risk prediction tool had less than optimal calibration; however, there was uncertainty in our results and excellent discrimination.

## Discussion

We assessed the performance of the International IgA risk prediction tool without race in a cohort from 2001 to 2016, which was primarily composed of the Hispanic and American Indian populations. Compared to the equation development cohort, our population had a higher baseline creatinine [median (IQR); 1.0 (0.8-1.4) vs. 2.14 (1.51 - 3.04)] and proteinuria [median (IQR); 1.2 (0.7 -2.2) vs. 2.7 (1.5-5.8)] around the time of IgA nephropathy diagnosis [5]. These patients were more likely to have higher segmental sclerosis (S1 77% vs. 88%) and tubular atrophy scores (T1 24.7 vs. 71% and T2 4.6 vs. 26%, respectively). Considering these findings, it is reasonable to assume that our cohort had a longer disease duration and severity. Indeed, the equation derivation population was younger when diagnosed with IgAN than the population in our study [Age(years); median (IQR); 35.6 (28.2 -45.4) vs. 38 (27-45)]. In our population, Hispanics were more likely to have segmental sclerosis on kidney biopsy (p 0.049) [Table 1]. For these reasons, the probability of achieving the primary outcome was higher in our cohort (68%) than in the equation development cohort (17.7%). All these findings may be explained by healthcare disparities faced by the Hispanic and American Indian residents of New Mexico. Race and ethnicity often provide insight into the socioeconomic and healthcare disparities in a population [14]. A review of the U.S. census 2017 data shows that in New Mexico, the poverty rates were higher for Hispanics (24.1 %) and American Indians (31.9%) than for Whites (17.7%) or Asians (10.5%) [15]. Furthermore, for each dollar earned by the White/Non-Hispanic residents of New Mexico, the American Indians and Hispanics earned approximately one-third less, highlighting the earnings disparities [16].

This study confirms the results of a previous study [10] looking at the frequency of IgAN in New Mexico in various races. US census data from 2010 showed that White/non-Hispanic, Hispanic, and American Indian shares of New Mexico’s population were 40.6, 46.4, and 8.5%, respectively [17]. The proportion of the Hispanics in our dataset (44%) matched their state population share; however, American Indians’ representation in our cohort was more than expected (26%) which may point towards a higher incidence of IgAN in the American Indian. This trend should be systematically evaluated in larger datasets.

A good prediction equation should be able to distinguish patients who are at low risk of achieving an outcome of interest from those who are at elevated risk. The international IgAN risk prediction tool did an excellent job identifying patients at a high risk of developing kidney failure early. The C-statistics, receiving operator curve, and the area under it (AUC) were close to what has been reported in the derivation and other validation cohorts (Table 2).

A well-calibrated risk equation’s predictions should match the observed risk. At 5 years, the equation overpredicted the risk for the patients to whom it assigned a risk of 20-50% and underpredicted the risk in patients to whom it assigned a higher risk (Figure 3); similar findings were also seen in the external validation of this equation in Chinese, Whites, and South Asians (Table 2)[6–8]. However, the equation calibration should be studied in a larger population to achieve more robust results.

**Figure 3:**
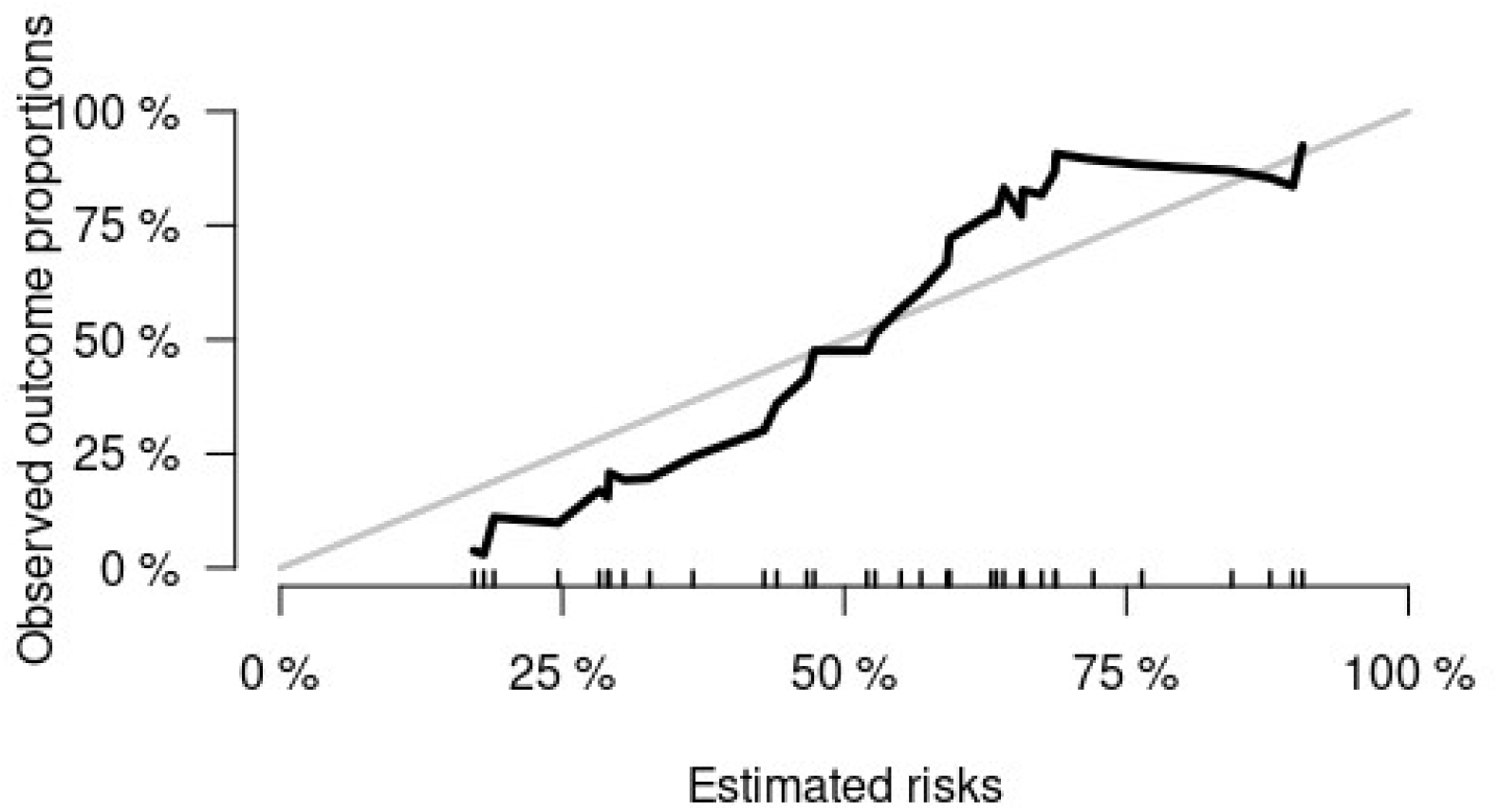
Calibration curve (Estimated risks plotted against Observed risk) at 5 years of the International IgAN risk prediction equation in the study population. Values below and above the 45 degrees line signify over and underprediction, respectively. The rug plot shows the distribution of the estimated risks.

Our study has weaknesses. First, our sample size is significantly smaller than the other populations in which this equation has been validated; therefore, there is uncertainty in our results, and a more extensive study is needed to assess the equation performance in American Indians and Hispanics. We used the bootstrapping technique for the metrics of C-statistics and IPA to obtain robust estimates and confidence intervals. Furthermore, despite of small sample size, the equation discrimination, and calibration performance match what has been reported in the other cohorts (Table 2). Second, our cohort had more severe disease than the equation development and the other cohorts in which it has been validated apart from the South Asian cohort [8], which could be the reason for the poor equation calibration seen in this study and underscores the need for an IgA risk-prediction equation developed in ethnically and racially diverse patients with advanced kidney disease.

Herein we provide the first report of IgA risk prediction tool performance in a population from the U.S. Southwest. We performed a rigorous chart review to ensure the accuracy of our data. We modeled the observed and predicted risks on a continuous scale which precluded the creation of arbitrary risk categories.

In summary, we showed that IgAN is diagnosed later in New Mexico, which may point towards economic and health care disparities. The international IgAN risk prediction tool without race can accurately and timely identify patients at a high risk of disease progression. In this cohort, the 5-year observed risk was lower in patients assigned a risk estimate of <50% of developing kidney failure and higher in patients assigned a risk estimate of >50%; these findings indicate over and under-prediction by the IgA International risk prediction tool, respectively. Our study suggests that the international IgAN risk prediction tool can be used to identify patients who are at a high risk of developing kidney failure; however, the calibration of the international IgAN risk prediction equation in our population was less than optimal and underscores the need of an IgAN risk prediction tool developed in a racially diverse population with more severe disease.

## Data Availability

Restrictions to the data apply; as per the University of New Mexico Institutional Review Board, an IRB modification with a data sharing agreement is required to provide data but can be arranged upon request.

## Disclosures

Saeed Kamran Shaffi:

Advisory board: Novartis

Christos Argyropoulos:

Consultant Agreements: Bayer, Baxter, Otsuka, Quanta

## Acknowledgments

The authors want to acknowledge the services of University of New Mexico Clinical and Translational Science Center (CTSC) who provided support for entering the data in the University of New Mexico Kidney biopsy registry. We would specifically like to thank Mr. Hugo A Vilchis and Greg Trejo on working on this project.

## Author Contribution

Saeed K Shaffi was responsible for funding acquisition for establishing the kidney biopsy registry, study conceptualization, project administration, data curation, and writing the original draft.

Edger Fischer reviewed the kidney biopsies and assigned a MEST-C score.

Christos Argyropolus devised study methodology and contributed to analysis.

Brent Wegner advised about project administration, validation and reviewing and editing of the manuscript.

