## Supplement 1 for "Validation of the International IgA risk prediction tool in American Indians and Hispanics"

Supplement 1 (S1): The risk prediction equation for the model without race

Predicted risk (time t) = 1 ‐ S0(t)Exp[LP]

Where:

LP = ‐0.320*[sqrt(eGFR)‐8.8] + 0.002*(MAP‐97) ‐ 0.035*[log(proteinuria) ‐ 0.09] + 0.004*[(MAP* log(proteinuria)) ‐ 8.73] + 0.201*M1 ‐ 0.035*E1 + 0.084*S1 + 0.700*T1 + 1.237*T2 + 0.101*T1*log(proteinuria) ‐ 0.321*T2*log(proteinuria) ‐ 0.017*(age‐38) + 0.118*RASB + 0.166*RASB* log(proteinuria) ‐ 0.266*immunosuppression

And

S0(t) = 1.0003754 ‐ 0.1131641*[(t+0.1)/100]2 + 0.0964763*[(t+0.1)/100]2 *log[(t+0.1)/100]
